# Nutritional status and its determinants among 0-5-year-old children in the district of Sirsa, Haryana, India

**DOI:** 10.1101/2025.08.30.25334626

**Authors:** Sanjay Kumar, Charu Khosla, Anup Kharde, Susmita Arya

**Affiliations:** School of Health Sciences, Chitkara University, Rajpura-140401, Punjab, India; School of Applied Sciences, Chitkara University, Rajpura-140401, Punjab, India; Pravara Institute of Medical Sciences (DU) Loni-413736, Maharashtra, India; Women & Child Development Department, Govt. of Haryana, Sirsa-125055, Haryana, India

**Keywords:** Under-nutrition, Young children, Risk factors, Interventions

## Abstract

**Background:** Researchers have hinted at the importance of balanced nutrition in infancy and childhood for physical development, cognitive function and robust immunity. Poor nutrition is known to produce several negative impacts on the physical and the mental health of the child. Assessing the actual condition of nutritional status of children pertaining to specific area often provide important insights for implementation of strategies necessary for prevention and control of under-nutrition. In this study, nutritional status and its determinants among 0-5-year-old children of Sirsa district of Haryana state were assessed.

**Methods:** A total of 300 children belonging to the age group 0-5 years, residing in the field practice area of Sirsa district of Haryana state, India, were included. A semi-structured questionnaire which was both pre-designed and pre-tested was used for data collection from the mothers of the children. The anthropometric measurements of the children were also taken.

**Results:** The prevalence rate of under-nutrition in this study was 7%. All of these children had sub-acute malnutrition (SAM). The anthropometric measurements (weight, height, head circumference, chest circumference, and mid-arm circumference) of under-nutrition children were significantly altered when compared with normal children. Bitot’s spot (57.1%) and rachitic changes (57.1%), followed by dry conjunctiva (47.6%), pedal oedema (42.9%), and corneal ulceration were common clinical signs/symptoms observed in SAM children. It was noted that a total of 46 (15.3%) children had moderate acute malnutrition (MAM). Upon ophthalmic examination, these children presented with dry conjunctiva.

**Conclusion:** The prevalence of under-nutrition in the form of sub-acute malnutrition and moderate acute malnutrition was 7% and 15.3% respectively. Under-nutrition was more common in girl child compared to male child. This study highlights the importance of conduction of regular screening program for early identification of extent and severity of under-nutrition in young children. Early identification of under-nutrition will facilitate the timely initiation of interventions necessary for normal growth and development of the child.

## Introduction

For any nation, the health of child is of paramount importance because ‘the children of today are citizen of tomorrow’. Health status in early ages of life impacts the physical and cognitive development of an individual, ultimately determining the future potential as an adult and citizen of the country, and therefore, it is very important to invest in the child’s health for a prosperous and healthy nation.^1^

From the prospective of life cycle, it is highly crucial to meet nutritional needs of the child in the first 1000 days of the life as during this period the requirement of nutrition is high due to rapid growth and development. It is the period when the physical development along with brain, metabolism and immune apparatus occurs.^2^ Therefore, any nutritional deficiency during this stage of life may deleteriously affect long term mental and physical wellbeing including delayed milestones, heightened susceptibility to infections, and increased risk of development of non-communicable diseases like diabetes, hypertension, obesity and cardiovascular diseases.^3^

Individual’s nutritional status is significantly influenced by three broad factors including food, health and care. These factors are interlinked and are known to differ from country to country and region to region within the country.^4^

Assessing the actual condition of nutritional status of children pertaining to specific area often provide important insights for implementation of strategies necessary for prevention and control of under-nutrition.^3, 4^ The present study was conducted with an aim to assess nutrition status and determinants among young children of Sirsa district of Haryana state.

## Material and Methods

Data collection for this study was conducted over a period of three months (April to June 2025) after obtaining ethical clearance from the Institutional Human Ethics Committee of Chitkara University, Rajpura, Punjab, India. The committee reviewed and approved the study protocol under approval number IHEC/DHR/CU/PB/24/323, ensuring compliance with institutional and ethical standards.

This was an observational study with cross-sectional design and included a total of 300 children belonging to age group 0-5 years residing in the field practice area of Sirsa district of Haryana state, India. The Sirsa district lies between 29 14 and 30 0 north latitude and 74 29 and 75 18 east longitudes, forming the extreme west corner of Haryana. This district is bounded by Faridkot and Bathinda districts of Punjab in the north and north east, Ganga Nagar district of Rajasthan in the west and south and Hisar district in the east.

A semi-structured questionnaire which was both pre-designed and pre-tested was used for data collection from the mothers of the children enrolled in the study. This questionnaire was filled by the investigator by interviewing technique. The questionnaire included questions based on the demography, breastfeeding practices, economic and socio-cultural factors. The anthropometric measurements of the children were also taken.

The collected data was entered in Microsoft Excel Spreadsheet and was appropriately coded. Statistical analysis was conducted by using SPSS (Statistical Package for Social Sciences) by IBM software program, version 25.5. As all tests were conducted at a 5% level of significance, an association was considered if the P value was <0.05.

## Results

Out of 300 participants, 153 (51%) were females whereas a total of 147 (49) participants were males. Although, the number of female participants was more compared to males, this difference was not statistically significant (Z test, *P*= 0.312). The mean age ± SD of the participants was 2.9±1.9 years. It ranged between 3 months to 5 years.

Age of a total of 29 participants was of less than 1 year whereas 171 participants were of age ≥ 1 year. The mean age ± SD of female participants was 2.4±1.4 years whereas the mean age ± SD of male participants was 2.7±1.2 years. There was no significant difference between the mean age of male and female participants (Mann Whitney U test, P>0.05).

A total of 192 (64%) participants were from rural parts whereas 108 (36%) participants were from urban areas. The number of participants were significantly higher from rural areas as compared to urban areas (Z test, *P<*.00001^**^). A total of 118 (39.3%) were staying in joint family, 108 (36%) were from nuclear family whereas 74 (24.7%) were staying with family having members of 3 generations.

When the socioeconomic status of family of the participants was assessed as per the BG Prasad Scale, it was noted that 93 (31%) participants were from socioeconomic (SE) Class V (lower class) and 79 (26.3%) belonged to SE Class IV (lower middle class). A total of 44 (14.7%) participants belonged to SE Class I (upper class). The participants from SE Class V were significant high as compared to other SE classes (Chi Square Test P<0.05^*^).

A total of 21 (7%) participants had under-nutrition, which included 9 boys and 12 girls. Therefore, the prevalence rate of malnutrition in this study was 7 %. All of these children had sub-acute malnutrition (SAM).

The mean age of the patients with under-nutrition was 2.8±1.2 years (Range 9 months-4.5 years). The mean ages of male and female participants were 2.9±1.7 years (Range 9 months to 4.5 years) and 2.6±0.7 years (Range 1.1 to 4 years). Although the mean age of male children was more than female children, this difference was not statistically significant (Mann Whitney U test, P>0.05). Out of 21 SAM children, 8 (38.1%) had partial vaccination whereas 13 (61.9%) reported to have complete vaccination.

The anthropometric measurements (weight, height, head circumference, chest circumference and mid arm circumference) of under-nutrition children were significantly altered when compared with normal children (Table 1).

**Table 1.**
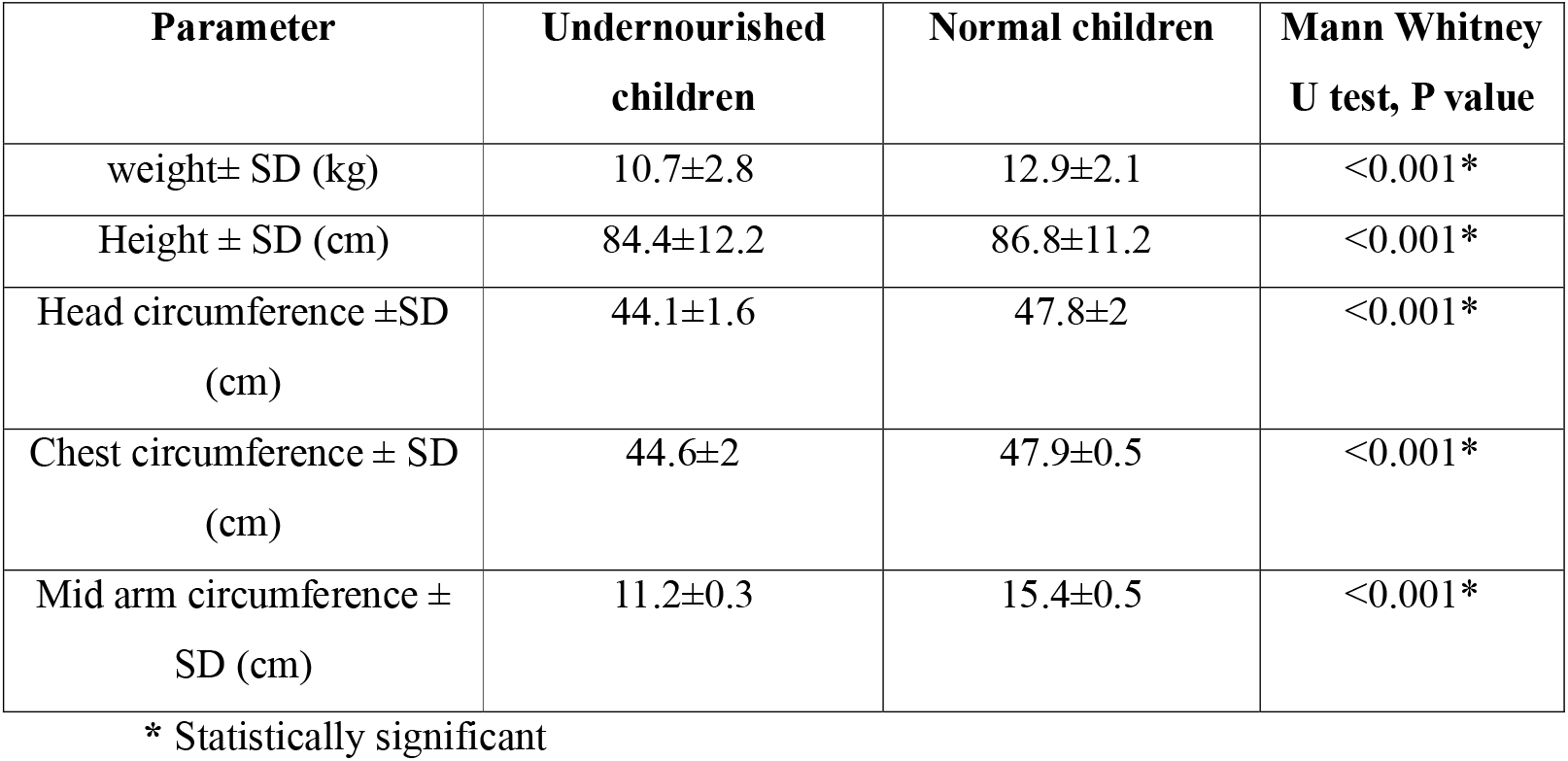
Comparison of anthropometric measurements of undernourished and normal children.

| Parameter | Undernourished children | Normal children | Mann Whitney U test, P value |
| --- | --- | --- | --- |
| weight± SD (kg) | 10.7±2.8 | 12.9±2.1 | <0.001* |
| Height ± SD (cm) | 84.4±12.2 | 86.8±11.2 | <0.001* |
| Head circumference ±SD (cm) | 44.1±1.6 | 47.8±2 | <0.001* |
| Chest circumference ± SD (cm) | 44.6±2 | 47.9±0.5 | <0.001* |
| Mid arm circumference ± SD (cm) | 11.2±0.3 | 15.4±0.5 | <0.001* |

\* Statistically significant

The clinical features of children with SAM are shown in table 2. Bitot’s spot (57.1%) and rachitic changes (57.1%) followed by dry conjunctiva (47.6%), pedal oedema (42.9%) and corneal ulceration were common clinical signs/symptoms observed in SAM children.

**Table 2.** Clinical features in under-nutrition children.

| **Clinical feature** | **N (%)** |
| --- | --- |
| Pedal oedema | 09 (42.9) |
| Dry conjunctiva | 10 (47.6) |
| Dry cornea | 01 (4.8) |
| Bitot's spot | 12 (57.1) |
| Corneal ulceration | 08 (38.1) |
| Xerophthalmia | 06 (28.6) |
| Rachitic changes | 12 (57.1) |
| Beading of ribs | 06 (28.6) |
| Pigeon chest | 07 (33.3) |

In this study when various parameters were studied in 300 children of age group 0-5 years belonging to Sirsa district of Haryana, it was noted that a total of 46 (15.3%) children had moderate acute malnutrition (MAM). Upon ophthalmic examination these children presented with dry conjunctiva.

The anthropometric measurements (weight, height, head circumference, chest circumference and mid arm circumference) of children with MAM is shown in table 3. When one way ANOVA test was used to compare these parameters in MAM, SAM and normal children, a significant alternation was observed (Table 3).

**Table 3.** Comparison of anthropometric parameters in MAM, SAM and normal children.

| **Parameter** | **MAM** | **SAM** | **Normal children** | **One way ANOVA, P value** |
| --- | --- | --- | --- | --- |
| weight± SD (kg) | 10.9±1.2 | 10.7±2.8 | 12.9±2.1 | <0.001* |
| Height ± SD (cm) | 83.8±9.8 | 84.4±12.2 | 86.8±11.2 | <0.001* |
| Head circumference<br>±SD (cm) | 44.7±1.8 | 44.1±1.6 | 47.8±2 | <0.001* |
| Chest circumference ±<br>SD (cm) | 45.1±1.4 | 44.6±2 | 47.9±0.5 | <0.001* |
| Mid arm<br>circumference ± SD<br>(cm) | 12.7±0.8 | 11.2±0.3 | 15.4±0.5 | <0.001* |

\* Statistically significant

## Discussion

Globally, malnutrition is one among the important risks for non-communicable diseases. Approximately one-third of world population have at least at least one form of malnutrition at any given point of the time resulting into stunting, wasting and micronutrient deficiencies. ^5^

Despite of several awareness programs, menace of child malnutrition is reported across the world. As per the data reported by the “United Nations Children (UNICEF)” and the “World Health Organization (WHO)”, 22.0% and 6.7% of children of the age less than 5□years were separately affected by stunting and wasting globally.^6^

In developing countries, annually, under-nutrition in children is a common problem and is associated with significant cause of morbidity and mortality.^7^ In Indian scenario, under-nutrition in children of less than five children is a major public health issue. In children of less than 5 years, underweight is reported in 39% to 75%, stunting is reported from 15.4% to 74% whereas wasting is evident in 10.6% to 42.3%. ^8^

The present study was in conducted in Sirsa district of Haryana, in which children belonging to age 0-5 years from different villages/cities like Baragudha, Odhan, Kalanwali, Chautala, Sirsa, Dabwali, Madho, Rania, Ellenabad and Chopta were included to determine the prevalence of undernutrition and its risk factors.

Out of these 300 participants 153 (51%) were females whereas a total of 147 (49) participants were males. Although the number of female participants were more than male participants, this difference was not statistically significant. Therefore, allocation of participants from both the gender was equal. In a similar conducted by Yadav et al.,(2016) from Haryana, male children were predominant.^9^ The mean age ± SD of the participants was 2.9±1.9 years. This finding was in consistent to other researchers.

In this study, out of 300 participants, a total of 21 (7%) participants had under-nutrition. Therefore, the prevalence rate of undernutrition in this study was 7 %. SAM is a life-threatening condition and is defined as ssevere wasting [weight-for-height Z score <−3 based on World Health Organization (WHO) reference standard] and/or the presence of nutritional oedema. ^10^

SAM needs urgent attention and appropriate management to reduce mortality and promote recovery. India has a high prevalence of SAM, representing a huge burden, and intriguingly, the recent National Family Health Survey-4 indicates a higher prevalence of severe wasting (7.5%) compared to the previous report (6.4%).^11^

Moderate acute malnutrition (MAM) was noted in a total of 46 (15.3%) children had moderate acute malnutrition (MAM). MAM affects 11% of children <5 y old worldwide and increases their risk for morbidity and mortality. It is assumed that successful treatment of MAM reduces these risks.^12^ Several risk factors are identified for malnutrition these include parental education, childhood illness, short birth interval, open defecation, type of weaning and complimentary food given.^13, 14^

In this study, lower socio-economic status of parents, illiteracy of parents and staying in nuclear family were profound risk factors associated with malnutrition. Study of these risk factors can be used for planning the diverse control measures.

## Conclusion

The prevalence of under-nutrition in the form of sub-acute malnutrition and moderate acute malnutrition was 7% and 15.3% respectively. Under-nutrition was more common in girl child compared to male child. This study highlights the importance of conduction of regular screening program for early identification of extent and severity of under-nutrition in young children. Early identification of under-nutrition will facilitate the timely initiation of interventions necessary for normal growth and development of the child.

## Data Availability

All data produced in the present study are available upon reasonable request to the authors.

## References

1. Chawla S, Gupta V, Singh A, Grover K, Panika RK, Kaushal P, Kumar A. Undernutrition and associated factors among children 1-5 years of age in rural area of Haryana, India: A community based cross-sectional study. J Family Med Prim Care. 2020; 9(8):4240–4246.

2. Likhar A, Patil MS. Importance of Maternal Nutrition in the First 1,000 Days of Life and Its Effects on Child Development: A Narrative Review. Cureus. 2022 Oct 8;14(10):e30083.

3. Wells JC, Dumith SC, Ekelund U, Reichert FF, Menezes AM, Victora CG, Hallal PC. Associations of intrauterine and postnatal weight and length gains with adolescent body composition: prospective birth cohort study from Brazil. Journal of Adolescent Health. 2012 Dec 1;51(6):S58–64.

4. Unicef. Improving child nutrition: the achievable imperative for global progress. New York: UNICEF. 2013 Dec;114.

5. Rakhshanda S, Barua L, Faruque M, Banik PC, Shawon RA, Rahman AKMF, Mashreky S. Malnutrition in all its forms and associated factors affecting the nutritional status of adult rural population in Bangladesh: results from a cross-sectional survey. BMJ Open. 2021;11(10):e051701.

6. Malnutrition Child: UNICEF & WHO. [Available from: https://data.unicef.org/topic/nutrition/malnutrition/.

7. Sulaiman AA, Bushara SO, Elmadhoun WM, Noor SK, Abdelkarim M, Aldeen IN, Osman MM, Almobarak AO, Awadalla H, Ahmed MH. Prevalence and determinants of undernutrition among children under 5-year-old in rural areas: A cross-sectional survey in North Sudan. J Family Med Prim Care. 2018;7(1):104–110.

8. Sahu SK, Kumar SG, Bhat BV, Premarajan KC, Sarkar S, Roy G, Joseph N. Malnutrition among under-five children in India and strategies for control. J Nat Sci Biol Med. 2015 Jan-Jun;6(1):18–23.

9. Yadav SS, Yadav ST, Mishra P, Mittal A, Kumar R, Singh J. An Epidemiological Study of malnutrition among under five children of rural and urban Haryana. J Clin Diagn Res. 2016; 10(2):LC07–10.

10. Kulkarni B, Mamidi RS. Nutrition rehabilitation of children with severe acute malnutrition: Revisiting studies undertaken by the National Institute of Nutrition. Indian J Med Res. 2019;150(2):139–152.

11. International Institute for Population Sciences. National Family Health Survey (NFHS-4) Mumbai: International Institute for Population Sciences; 2016.

12. Chang CY, Trehan I, Wang RJ, Thakwalakwa C, Maleta K, Deitchler M, Manary MJ. Children successfully treated for moderate acute malnutrition remain at risk for malnutrition and death in the subsequent year after recovery. The Journal of nutrition. 2013;143(2):215–20.

13. Jeyaseelan V, Jeyaseelan L, Yadav B. Incidence of, and risk factors for, malnutrition among children aged 5–7 years in South India. Journal of Biosocial Science. 2016; 48(3):289–305.

14. Chakravorty S, Manisha. Data analysis of malnutrition in India: a review of numerous factors. Int J Community Med Public Health 2023; 10(7):2629–36.

